# Disparities in stroke incidence over time by sex and age in Latin America and the Caribbean region: a systematic review and meta-analysis

**DOI:** 10.1101/2023.02.23.23286066

**Authors:** Marilaura Nuñez, Carlos Delfino, Claudia Asenjo-Lobos, Andrea Schilling, Pablo Lavados, Craig S. Anderson, Paula Muñoz Venturelli

## Abstract

**Background:** High-income countries studies show unfavorable trends in stroke incidence (SI) in younger populations. We aimed to estimate temporal change in SI disaggregated by age and sex in Latin America and the Caribbean region (LAC).

**Methods:** A search strategy was used in MEDLINE, WOS, and LILACS databases from 1997 to 2021, including prospective observational studies with age and sex-disaggregated data of first-ever stroke (FES) incidence. Risk of bias was assessed with The Joanna Briggs Institute’s guide. The main outcomes were incidence rate ratio (IRR) and relative temporal trend ratio (RTTR) of SI, comparing time periods ≥2010 with <2010. Pooled RTTR (pRTTR) only considering studies with two periods in the same population were calculated by random-effects meta-analysis.

**Results:** From 9,242 records identified, six studies were selected including 4,483 FES in 4,101,084 individuals. Crude IRR in younger subjects (<55 years) comparing ≥2010:<2010 periods showed an increase in SI in the last decade (IRR 1.37;95%CI 1.23-1.50), in contrast to a decrease in older people during the same period (IRR 0.83; 95%CI 0.76–0.89). Overall RTTR (<55:≥55 years) was 1.65 (95CI% 1.50-1.80), with higher increase in young women (pRTTR 3.08; 95%CI 1.18-4.97; p for heterogeneity <0.001).

**Conclusions:** An unfavorable change in SI in young people - especially in women - was detected in the last decade in LAC. Further investigation of the explanatory variables is required to ameliorate stroke prevention and inform local decision-makers.

**Registration of protocol:** CRD42022332563 (PROSPERO).

## Introduction

Stroke is one of the most important causes of combined death and disability, worldwide^1^. Efforts to reduce the incidence of stroke and to guide health policies have been implemented in various countries. Stroke prevention has focused on reducing systolic arterial hypertension, high fasting plasma glucose and cholesterol, high body index mass, smoking, and other factors^1–3^. However, these risk factors are becoming increasingly prevalent in young people^3,4^.

The Global Burden Disease has reported that from 1990 to 2019, there has been an increase in the burden of stroke in low- and upper-middle-income countries. Moreover, an increase in the incidence of stroke in those under 70 years of age, with a substantial increase in the last decade (2010-2019)^5^.

Recently, studies in high-income countries reported divergent temporal trends in stroke incidence at age 55 years and older. Younger people (<55 years) had a less favorable incidence of stroke in the last decade compared with older people in the same period (≥55 years)1. Unique features of the burden of stroke in Latin America and the Caribbean (LAC) region are that rates are high compared to averages, worldwide^6^ and there is no assessment of age and sex disparities that have been noted elsewhere. Since local data can guide future policies, we aimed to estimate temporal trends in stroke incidence disaggregated by age and sex in LAC.

## Methods

A systematic review is reported according to Metanalysis of Observational Studies in Epidemiology (MOOSE) and Preferred Reporting Items for Systematic reviews and Meta-Analyses (PRISMA) guidelines^7^ (Fig. S1). The protocol was registered in PROSPERO (CRD42022332563).

### Selection criteria and search strategy

Prospective observational studies in LAC region from 1997 to 2021 reporting age and sex-disaggregated data on first-ever stroke (FES) incidence were included. FES reporting had to be supported by clinical and/or imaging diagnosis. Only studies published in English, Portuguese and Spanish were considered. Retrospective studies, reviews, commentaries, and editorials were excluded.

The search strategy followed the same parameters of our previous publication^8^. A modified search strategy of the Cochrane Stroke Group filtered by terms for Latin American and the Caribbean was used to identify studies that met the selection criteria. The search strategy was conducted between January 1, 1997 and December 31, 2021 from MEDLINE (Ovid), Lilacs and Science Citation Index Expanded (SCI—EXPANDED), Social Sciences Citation Index and Arts, and Humanities Citation Index within ISI Web of Science (WoS) (from inception to May 27, 2022) (Fig. S2-S4).

### Selection process and data extraction

Two independent reviewers (CD and MN), prior to the duplicate elimination process in EndNote 20, analyzed and selected all relevant abstracts or titles through the Joanna Briggs Institute software (http://www.jbisumari.org/)^9^. In case of disagreement a third reviewer was necessary to reach a decision.

A standardized form was made with the data of the article to be extracted: authors, periods, city, country of study and annual incidence rate of FES. The annual incidence rate of FES per 100,000 people, disaggregated for age and sex for the LAC region between January 1, 1997 and December 31, 2021 was the main extracted result.

### Studies quality and risk of bias assessment

Two independent reviewers assessed data quality and study design. A third reviewer was consulted in case of disagreement (Table S1). Risk of bias assessment was performed using the Joanna Briggs Institute’s (JBI) checklist for a cohort studies^9^.

### Data synthesis and analysis

Disaggregated data of incidence rate (IR) were extracted from each article by sex and age (men: women, and age groups of <55 vs. ≥55 years and <45 vs. ≥45 years). The cut-off age of 55 was selected because it is the youngest age criterion used in previous epidemiological studies^1^. When enough data were available, a cutoff age of 45 years was used, given current concerns regarding stroke in the young. Incidence rate ratio (IRR) comparing two time periods (incidence rate ≥2010: incidence rate <2010) by sex and age group were calculated. A cut-off in year 2010 was considered because several initiatives have been implemented to improve stroke care in LAC countries after this time point^10^. The overall relative temporal trend ratio (RTTR) comparing age groups in the overall population and by sex groups were determined (IRR<55:IRR≥55 and IRR<45:IRR≥45 years)1. Pooled RTTRs (pRTTR) including only studies with 2 time periods with the same population were calculated by random-effects meta-analysis. When two periods in the same population were compared in the pRTTR, the time difference between the earliest and the latest time period was 10.6 years (SD 1.8)^1^. A subgroup analysis by sex was performed. The heterogeneities of the pRTTRs were calculated using Cochrane’s Q test. All of outcomes were processed with a 95% confidence interval (CI). STATA version 17.1 was used for statistical analysis.

## Results

Of 9,242 identified records, six prospective observational studies (nine periods) in LAC between 1997 and 2021 fulfilled the eligibility criteria^2,11–15^ (Table 1). A total of 4,483 FES in 4,101,084 individuals were considered. The Prisma flow diagram is available in supplementary material (Fig. S5).

**Table 1.**
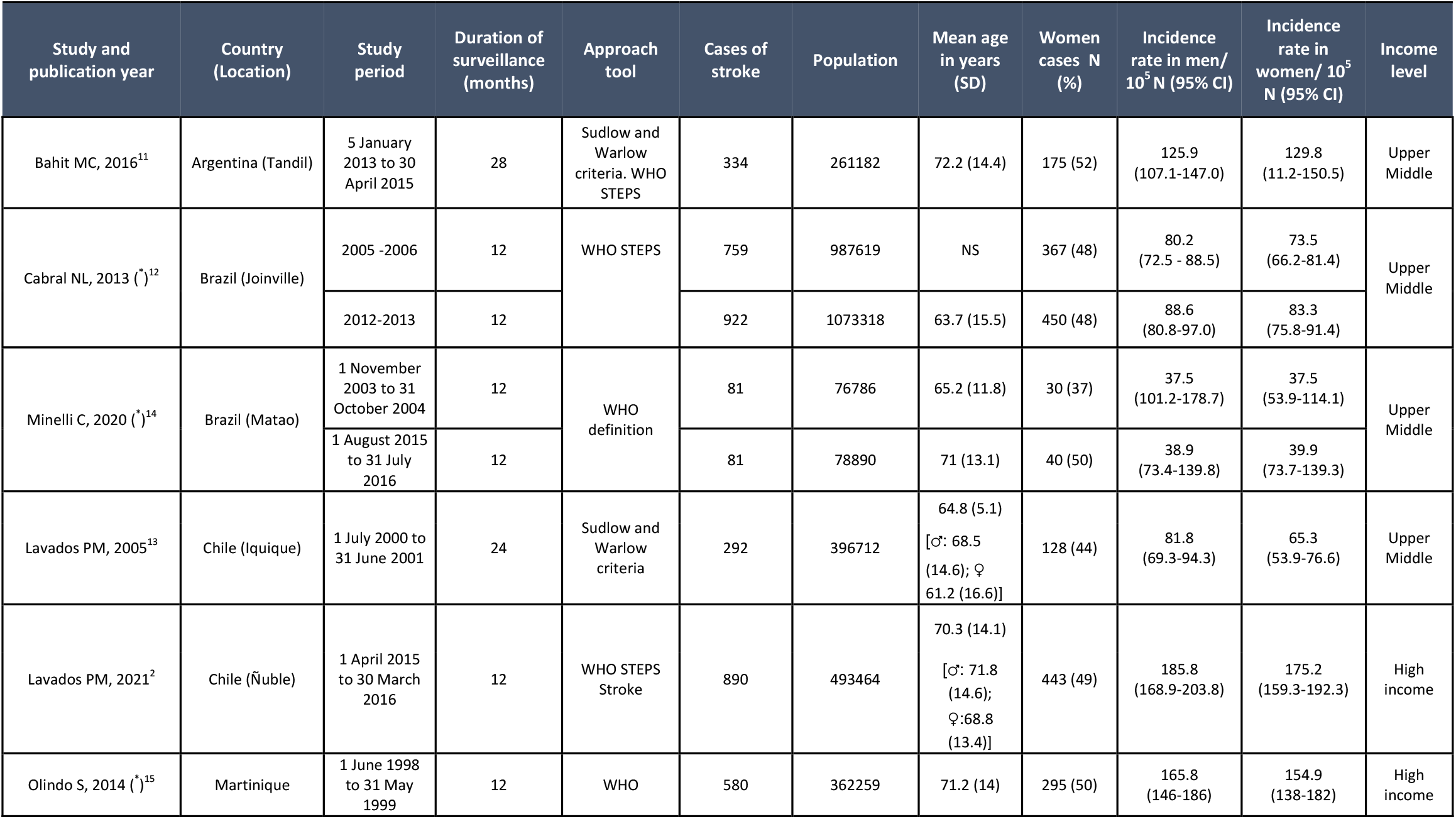
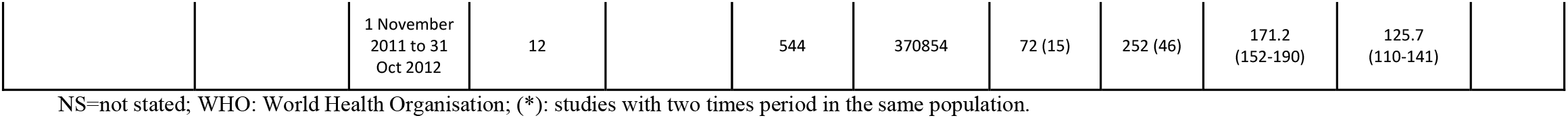
Summary of the data from studies in Latin America and the Caribbean

| Study and publication year | Country (Location) | Study period | Duration of surveillance (months) | Approach tool | Cases of stroke | Population | Mean age in years (SD) | Women cases N (%) | Incidence rate in men/ 10<sup>5</sup> N (95% CI) | Incidence rate in women/ 10<sup>5</sup> N (95% CI) | Income level |
| --- | --- | --- | --- | --- | --- | --- | --- | --- | --- | --- | --- |
| Bahit MC, 2016<sup>11</sup> | Argentina (Tandil) | 5 January 2013 to 30 April 2015 | 28 | Sudlow and Warlow criteria. WHO STEPS | 334 | 261182 | 72.2 (14.4) | 175 (52) | 125.9 (107.1-147.0) | 129.8 (11.2-150.5) | Upper Middle |
| Cabral NL, 2013 (*)<sup>12</sup> | Brazil (Joinville) | 2005 -2006 | 12 | WHO STEPS | 759 | 987619 | NS | 367 (48) | 80.2 (72.5 - 88.5) | 73.5 (66.2-81.4) | Upper Middle |
|  |  | 2012-2013 | 12 |  | 922 | 1073318 | 63.7 (15.5) | 450 (48) | 88.6 (80.8-97.0) | 83.3 (75.8-91.4) |  |
| Minelli C, 2020 (*)<sup>14</sup> | Brazil (Matao) | 1 November 2003 to 31 October 2004 | 12 | WHO definition | 81 | 76786 | 65.2 (11.8) | 30 (37) | 37.5 (101.2-178.7) | 37.5 (53.9-114.1) | Upper Middle |
|  |  | 1 August 2015 to 31 July 2016 | 12 |  | 81 | 78890 | 71 (13.1) | 40 (50) | 38.9 (73.4-139.8) | 39.9 (73.7-139.3) |  |
| Lavados PM, 2005<sup>13</sup> | Chile (Iquique) | 1 July 2000 to 31 June 2001 | 24 | Sudlow and Warlow criteria | 292 | 396712 | 64.8 (5.1)<br>[♂: 68.5 (14.6); ♀: 61.2 (16.6)] | 128 (44) | 81.8 (69.3-94.3) | 65.3 (53.9-76.6) | Upper Middle |
| Lavados PM, 2021<sup>2</sup> | Chile (Ñuble) | 1 April 2015 to 30 March 2016 | 12 | WHO STEPS Stroke | 890 | 493464 | 70.3 (14.1)<br>[♂: 71.8 (14.6); ♀: 68.8 (13.4)] | 443 (49) | 185.8 (168.9-203.8) | 175.2 (159.3-192.3) | High income |
| Olindo S, 2014 (*)<sup>15</sup> | Martinique | 1 June 1998 to 31 May 1999 | 12 | WHO | 580 | 362259 | 71.2 (14) | 295 (50) | 165.8 (146-186) | 154.9 (138-182) | High income |
|  |  | 1 November<br>2011 to 31<br>Oct 2012 | 12 |  | 544 | 370854 | 72 (15) | 252 (46) | 171.2<br>(152-190) | 125.7<br>(110-141) |  |

NS=not stated; WHO: World Health Organisation; (\*): studies with two times period in the same population.

### Incidence Rate Ratio

The IR in men was higher than in women with an IRR 1.12 (95%CI 1.04-1.21 [I^2^ =38.3%, *P*=0.11]). (Figure 1A).

**Fig. 1.**
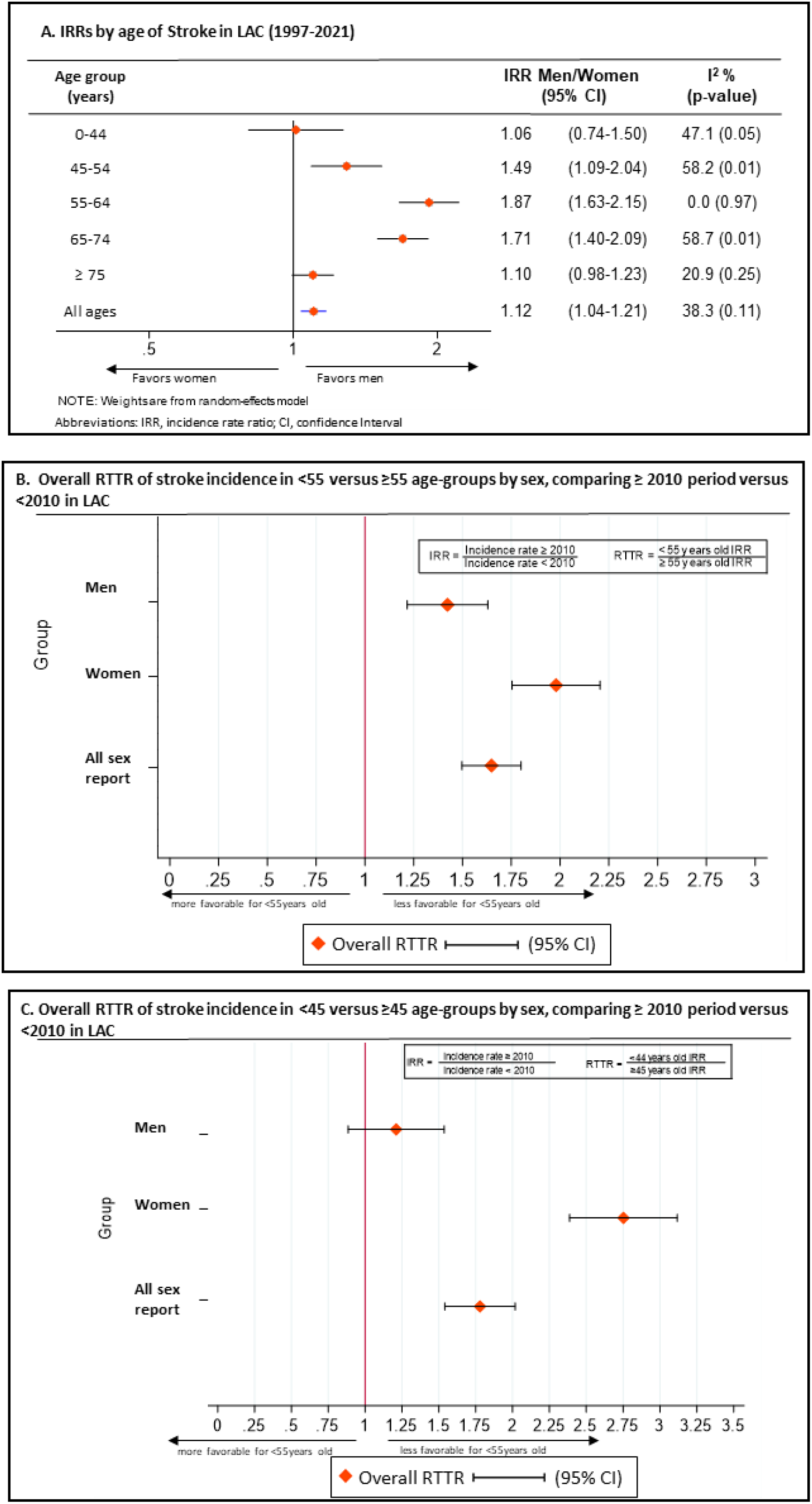
Incidence Rate Ratio (IRR) by age and overall Relative Temporal Trend Ratios (RTTR) of stroke in Latin America and the Caribbean (LAC)

Crude IRR for stroke by age was compared between 2 periods of time (≥2010 and <2010). Those aged less than 55 years were found to have a higher IR in the latest period (≥2010) compared to the previous period (IRR 1.37, 95%CI 1.23-1.50). In contrast, the IR of the older group (≥55 years) decreased in the same periods (IRR 0.83, 95%CI 0.76–0.89).

Crude IRR for stroke by age was analyzed using a cutoff point of <45 years old, and also showed a worse trend in the last decade for those <45 years (≥2010:<2010; IRR 1.59, 95%CI 1.36-1.82). Crude IRR for stroke in ≥45 years age-group showed a reduction in stroke incidence in the last decade (≥2010:<2010; IRR 0.89, 95%CI 0.83-0.95) that was similar to the ≥55 years age-group.

### Relative Temporal Trend Ratio

When comparing incidence period ≥2010 to <2010 between age groups, the overall RTTR for stroke incidence was less favorable in those <55 years old (IRR <55 vs. IRR ≥55 years; RTTR 1.65, 95%CI 1.50-1.80) (Figure 1B). Overall, women experienced a higher increase in stroke incidence (<55 vs. ≥55 years; women RTTR 1.98, 95%CI 1.75-2.21) compared to men (<55 vs. ≥55 years; men RTTR 1.42, 95%CI 1.22-1.63).

Overall RTTR was less favorable in <45 than ≥45 years age-groups comparing incidence periods of ≥2010 to <2010 (IRR <45 vs. IRR ≥45 years; RTTR 1.77, 95%IC 1.54-2.01) (Figure 1C). These findings had similar RTTR to the first cut-off point (<55 vs. ≥55 years old), but when disaggregated by sex, the magnitude of RTTR was higher in younger women compared to younger men (women RTTR 2.75, 95%CI 2.38–3.11 vs. men RTTR 1.21, 95%CI 0.88-1.53).

### Pooled Relative Temporal Trend Ratio

When only studies reporting two periods in the same population with the <55 years age cutoff point were included, the pRTTR was similar to overall RTTR (<55 vs. ≥55 years; pRTTR 1.77, 95%CI 1.59-1.95; *P* for heterogeneity = 0.59) (Figure 2A). When disaggregated by sex, pRTTR showed similar outcomes in both sexes to those of the RTTR analysis, (women pRTTR 2.23; 95CI%1.41-3.04; p for heterogeneity 0.004 and men pRTTR 1.48; 95CI% 1.23-73; p for heterogeneity 0.72).

**Fig. 2.**
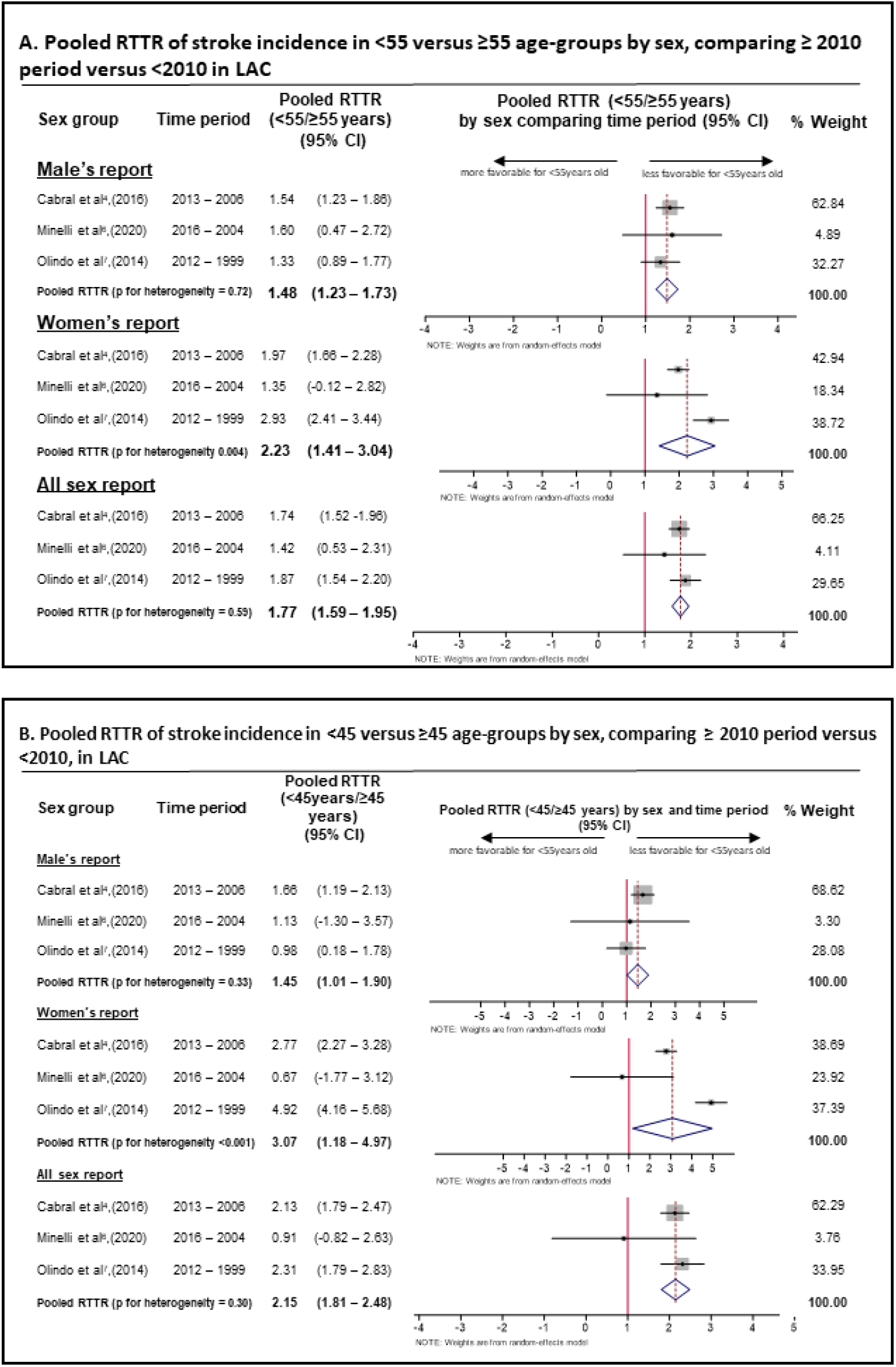
Pooled RTTR of stroke incidence by age – groups

Using <45 years cutoff point, the pRTTR (<45 vs. ≥45 years; pRTTR 2.15, 95%CI 1.81-2.48; *P* for heterogeneity = 0.30) was also comparable to overall RTTR (Figure 2B). Analysis by sex maintains the magnitude of RTTR at <45 cutoff point (women pRTTR 3.08; 95CI% 1.18-4.97; p for heterogeneity <0.001 and men pRTTR 1.45; 95CI% 1.01-1.90; p for heterogeneity 0.33)

Subgroup funnel plots showed symmetrical representation and all Egger’s tests were ≥0.1. Both tests were applied to assess the risk of bias (Figure S6).

## Discussion

Our systematic review and meta-analysis has shown an unfavorable temporal trend in stroke incidence in LAC for younger people, particularly in young women. This contrasts with a global decline in stroke incidence during 2010-2019 compared to 2000-2009, which has been driven by older populations^1,5^.

The divergence of temporal trends in stroke incidence, with an increase in those <55 years and a decrease in people ≥55 years in the most recent decade (≥2010) compared with the earlier period (<2010), is consistent with the results of a recent prospective population-based incidence study in England^16^, and other similar data from high income countries in the 21^st^ century^1^. In younger people, unfavorable outcomes may be associated with increases in conventional vascular risk factors, such as hypertension, obesity, sedentary lifestyle, unhealthy diets, dyslipidemia, diabetes mellitus, and smoking^2,13,15^. Also, the emergence of new risk factors such as air pollution, long working hours, and stress, but also better patient awareness and access to acute stroke diagnosis may also have a role in these changes^1,17^. Hormonal factors could be specifically relevant to younger women^12,14,18^.

The strengths of this systematic review include the availability of population-based studies, most of which met the suggested ‘standard or advanced criteria for ideal stroke incidence studies’ and covered a long period^1,19^, with age and sex disaggregated data available. However, there are only a limited number of published reports from few countries, so our results should be interpreted with caution due to possible selection bias, the potential for unmeasured confounders, the lack of information on stroke severity/type and prevention strategies; and heterogeneity from the use of different methodology between older studies compared to more recent ones. In addition, even though our data were derived from varied populations (likely causing clinical heterogeneity) which may differ from other LAC countries where stroke incidence data were not available, particularly those in lower-income areas.

The unfavorable temporal trend in stroke incidence in LAC for younger populations, particularly in young women, highlighted in our study warrants further research to understand explanatory factors in order to guide future health policies and to prevent further increases in the disease burden.

## Registration of protocol

CRD42022332563 (PROSPERO).

## Data Availability

All data will be provided by the corresponding author upon reasonable request.

## Abbreviations

IR: Incidence rate
IRR: Incidence rate ratio
FES: First-ever stroke
LAC: Latin American and Caribbean
pRTTR: pooled relative temporal trend ratio
RTTR: Relative temporal trend ratio
SI: Stroke incidence

## Acknowledgment

None.

