## Supplemental Material for "Disparities in stroke incidence over time by sex and age in Latin America and the Caribbean region: a systematic review and meta-analysis"

**eFigure 1.** PRISMA Checklist

**eFigure 2.** WOS search algorithm

**eFigure 3.** MEDLINE search algorithm

**eFigure 4.** LILACS search algorithm

**eFigure 5**. PRISMA Flow Diagram

**eFigure 6**. Funnel plots and Egger´s test used to assess risk of bias by subgroup

**eTable 1.** JBI´s critical appraisal for included studies

.

**eFigure 1. PRISMA Checklist**
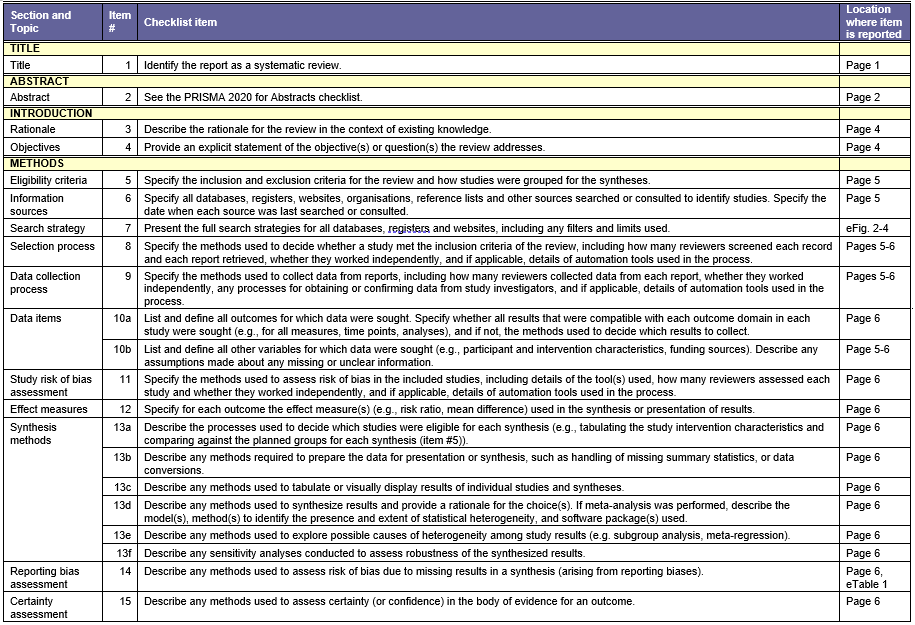

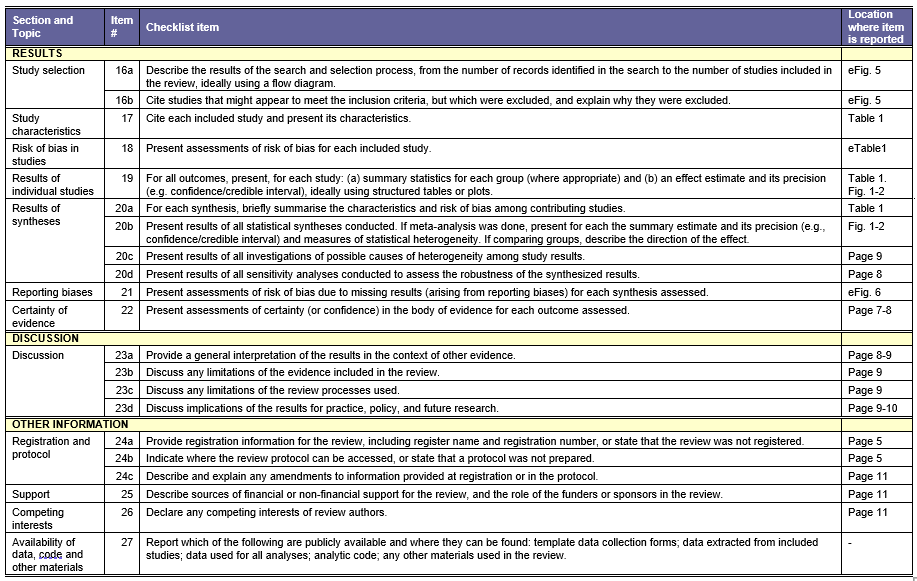

**eFigure 2**
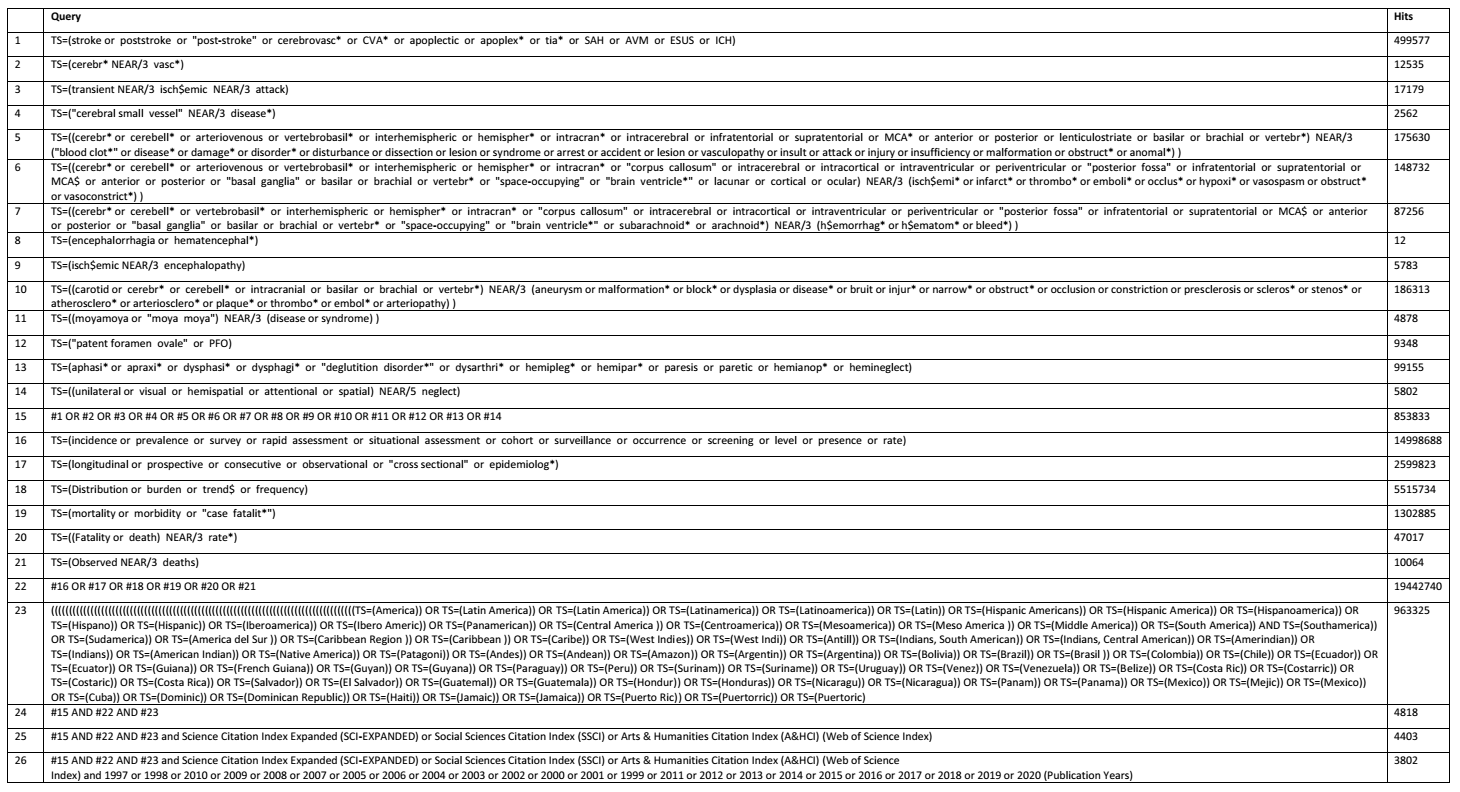
**. WOS search algorithm**
**eFigure 3. MEDLINE search algorithm**

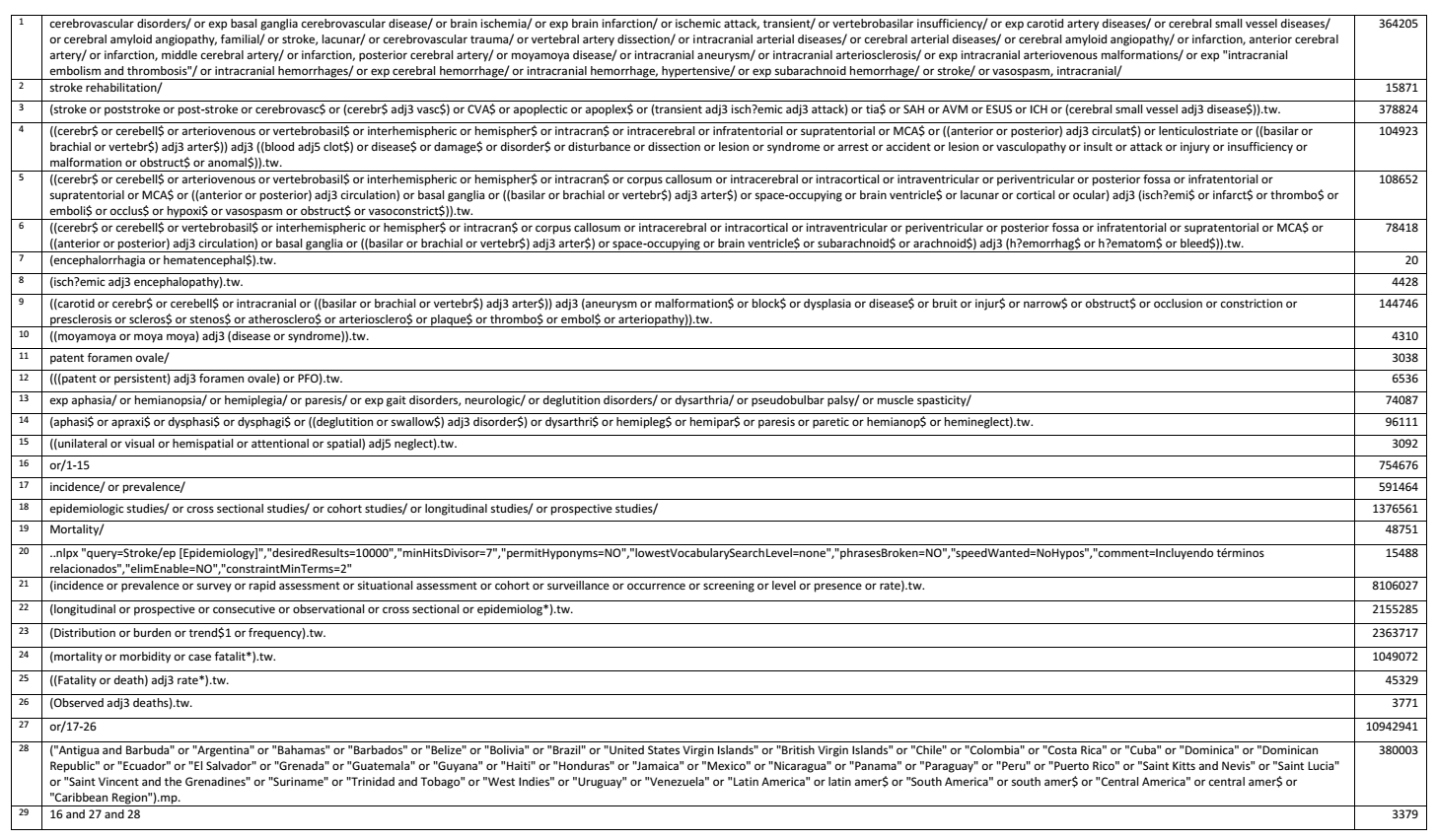

**eFigure 4. LILACS search algorithm**

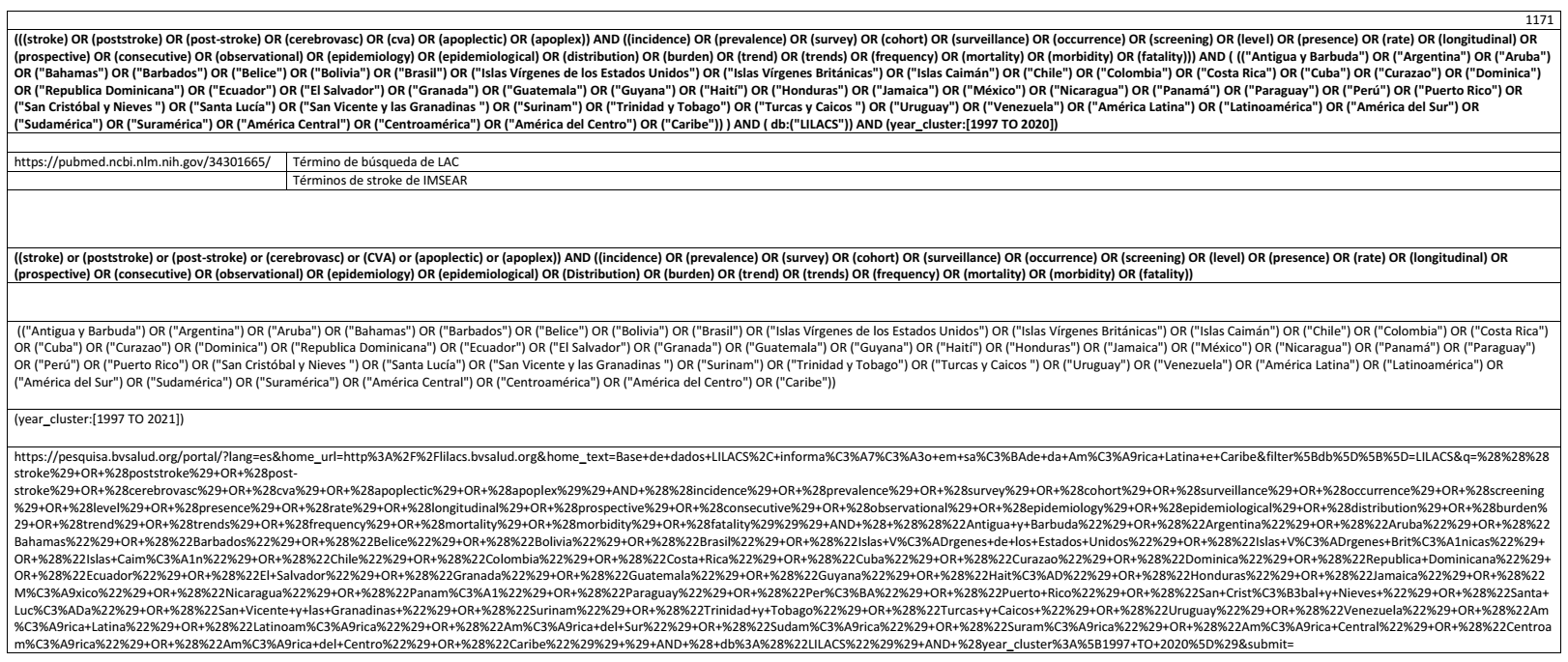

**eFigure 5. PRISMA Flow Diagram**

**Identification**

**Included**

Records screened

(n = 7,301)

Records excluded

(n = 7,185)

Studies sought for retrieval

(n = 116)

Studies assessed for eligibility

(n = 116)

Reports excluded (n = 110):

Excluded study design (n = 13)

Conference/poster abstract (n = 17)

Editorial commentary (n = 5)

Letter to editor (n = 3)

Review (n = 3)

Duplicate data (n = 13)

Not interested outcome (n = 45)

Follow-up outside the period (n = 5)

Unable to obtain (n = 6)

Studies included in review

(n = 6)

Records identified from:

WOS database

(n = 4,193)

Records identified from:

MEDLINE database

(n = 3,819)

Records identified from:

LILACS database

(n = 1,230)

Records removed before screening:

Duplicate records removed (n = 1,941)

**Screening**

**eFigure 6. Funnel plots and Egger´s test used to assess risk of bias by subgroup**

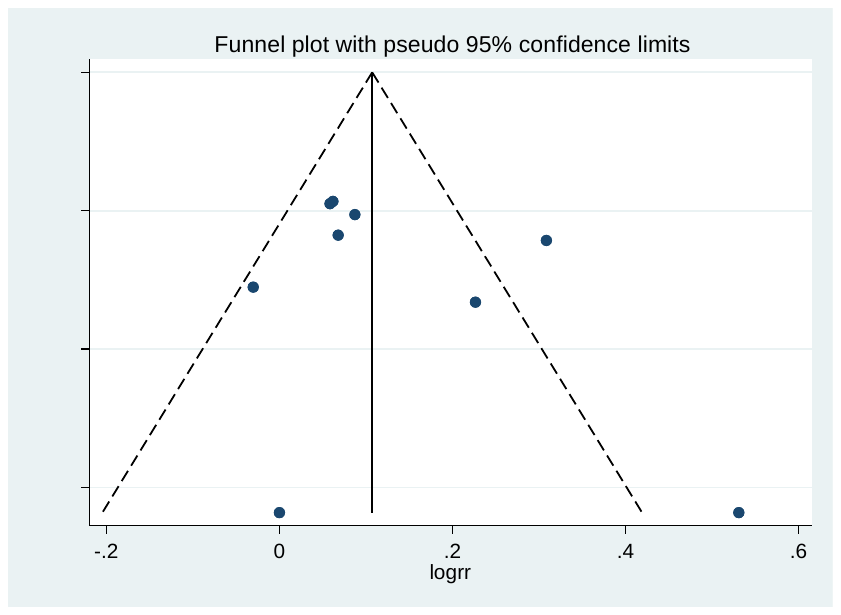

Egger's test for small-study effects (*P* = 0.346 )

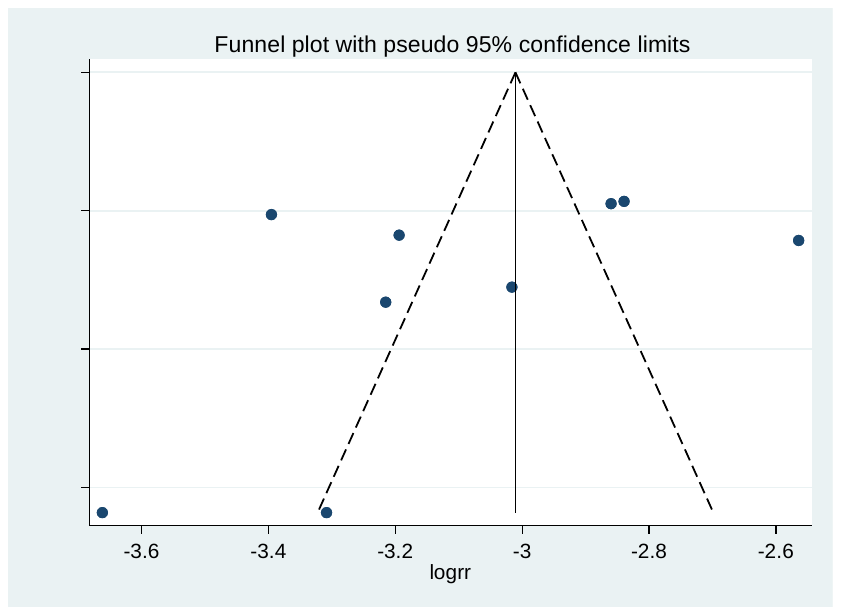

Egger's test for small-study effects (*P* = 0.339 )

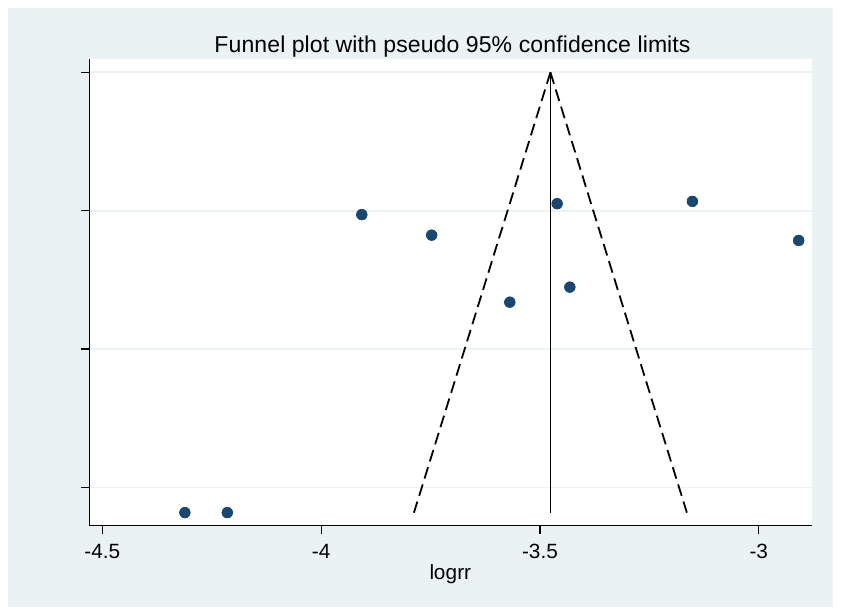

Egger's test for small-study effects (*P* = 0.296 )

**A. Analysis by sex subgroup**

**B. Analysis by cutoff point of <55 years**

**C. Analysis by cutoff point of <45 years**

**eTable 1. JBI´s critical appraisal for included studies**

| **Incidence and case-fatality studies** | | | | | | | | | | | | |  |
| --- | --- | --- | --- | --- | --- | --- | --- | --- | --- | --- | --- | --- | --- |
| **ID** | **Citation** | **Q1** | **Q2** | **Q3** | **Q4** | **Q5** | **Q6** | **Q7** | **Q8** | **Q9** | **Q10** | **Q11** | **Total** |
| 1 | Bahit MC, et al. 2016. | Y | Y | Y | Y | Y | Y | Y | Y | Y | Y | Y | 11/11 |
| 2 | Cabral NL, et al. 2016. | Y | Y | Y | Y | Y | Y | Y | Y | Y | Y | Y | 11/11 |
| 3 | Lavados PM, et al. 2005 | Y | Y | Y | Y | Y | Y | Y | Y | Y | Y | Y | 11/11 |
| 4 | Lavados PM, et al. 2021 | Y | Y | Y | Y | Y | Y | Y | Y | Y | Y | Y | 11/11 |
| 5 | Minelli C, et al. 2020. | Y | Y | Y | Y | Y | Y | Y | Y | Y | Y | Y | 11/11 |
| 6 | Olindo S, et al. 2014 | Y | Y | Y | Y | Y | U | U | Y | Y | Y | N | 9/11 |
| Legend: ID, identification number; Q, question; Y, yes; N, no | | | | | | | | | | | | | |
